# Predicting Cerebrospinal Fluid Alpha-Synuclein Seed Amplification Assay Status from Demographics and Clinical Data

**DOI:** 10.1101/2024.08.07.24311578

**Authors:** Charles S. Venuto, Konnor Herbst, Lana M. Chahine, Karl Kieburtz

**Author notes:** Correspondence to: Charles S. Venuto, Center for Health + Technology, University of Rochester, 265 Crittenden Blvd., Rochester, NY 14642.

## Abstract

**Objective:** To develop and externally validate models to predict probabilities of alpha-synuclein (a-syn) positive or negative status in vivo in a mixture of people with and without Parkinson’s disease (PD) using easily accessible clinical predictors.

**Methods:** Uni- and multi-variable logistic regression models were developed in a cohort of participants from the Parkinson Progression Marker Initiative (PPMI) study to predict cerebrospinal fluid (CSF) a-syn status as measured by seeding amplification assay (SAA). Models were externally validated in a cohort of participants from the Systemic Synuclein Sampling Study (S4) that had also measured CSF a-syn status using SAA.

**Results:** The PPMI model training/testing cohort consisted of 1260 participants, of which 76% had manifest PD with a mean (± standard deviation) disease duration of 1.2 (±1.6) years. Overall, 68.7% of the overall PPMI cohort (and 88.0% with PD of those with manifest PD) had positive CSF a-syn SAA status results. Variables from the full multivariable model to predict CSF a-syn SAA status included age- and sex-specific University of Pennsylvania Smell Identification Test (UPSIT) percentile values, sex, self-reported presence of constipation problems, leucine-rich repeat kinase 2 (*LRRK2*) genetic status and pathogenic variant, and *GBA* status. Internal performance of the model on PPMI data to predict CSF a-syn SAA status had an area under the receiver operating characteristic curve (AUROC) of 0.920, and sensitivity/specificity of 0.881/0.845. When this model was applied to the external S4 cohort, which included 71 participants (70.4% with manifest PD for a mean 5.1 (±4.8) years), it performed well, achieving an AUROC of 0.976, and sensitivity/specificity of 0.958/0.870. Models using only UPSIT percentile performed similarly well upon internal and external testing.

**Conclusion:** Data-driven models using non-invasive clinical features can accurately predict CSF a-syn SAA positive and negative status in cohorts enriched for people living with PD. Scores from the UPSIT were highly significant in predicting a-syn SAA status.

## INTRODUCTION

The introduction of the α-synuclein (a-syn) cerebrospinal fluid (CSF) seeding amplification assay (SAA) offers a promising approach for the diagnosis of Parkinson’s disease (PD) and other synucleinopathies. This method is notable for its high sensitivity and specificity in distinguishing between people with and without PD.^1,2^ Additionally, there is an emerging emphasis on a-syn SAA positive results to redefine and stage PD biologically.^3-5^ These approaches promote a shift away from solely relying on symptom-based diagnostics, and instead focus towards a framework that integrates biological markers of a-syn aggregation, thus enhancing the potential for earlier detection and intervention.

However, the implementation of CSF a-syn SAA is not without its challenges. In particular, the assay requires a spinal tap for the collection of CSF samples, an invasive procedure that may limit its routine clinical use. Therefore, efforts are ongoing to extend SAA applicability to other more accessible fluids and tissues. For instance, detection of a-syn in skin biopsy samples has shown good performance similar to those using CSF samples for differentiating PD from controls.^6-10^ However, fewer studies have evaluated skin biopsies, and technical aspects such as number of biopsies, sampling locations, and sample processing still need to be optimized. Additional biospecimens, such as saliva, blood, submandibular gland, and colonic mucosa are still under development and are not as readily available.^11-14^ Therefore, at present, CSF remains the leading biomatrix for the detection of a-syn using SAA.

Intriguingly, in the largest study to date that measured CSF a-syn via SAA in a predominantly PD population, the proportion of individuals yielding a positive CSF a-syn SAA result was not uniform. For instance, only 67.5% of individuals with PD carrying genetic mutations in the leucine-rich repeat kinase 2 (*LRRK2*) gene were positive for a-syn CSF SAA compared to 93.3% of sporadic PD individuals. For PD participants without hyposmia, the sensitivity of the a-syn SAA assay was only 63.0%.^15^ This variation when stratified by genetic and clinical characteristics, points to a nuanced relationship between select clinical features and the disease pathology. We aimed to leverage this variation by developing data-driven models that can predict CSF a-syn SAA status probabilities based on simple, non-invasively collected assessments.

## METHODS

### Study Data

Model training data came from the Parkinson’s Progression Marker Initiative (PPMI) study cohort. PPMI is a longitudinal observational study that is recruiting participants from outpatient neurology clinics at academic centers in North America and Europe (registered at ClinicalTrials.gov NCT01141023).^16^ The objective of the PPMI study is to identify clinical and biological markers of PD risk, onset, and progression. Enrollment began in 2010 and is ongoing. Participants in PPMI were included in one of the following cohorts: untreated sporadic PD; PD with presence of pathogenic genetic variants (*LRRK2*, *GBA*, *SNCA*, *Parkin*, *PINK1*); at-risk non-manifest PD based on presence of rapid eye movement (REM) sleep behavior disorder (RBD), hyposmia with dopamine transporter (DAT) deficit measured using single photon emission computed tomography (SPECT), or presence of a genetic risk variant; ‘healthy controls’ who have no neurologic disorder and no first-degree relative with PD; and, people with parkinsonism but with DAT SPECT scans without evidence of dopamine deficiency (SWEDD). The diagnosis for each cohort was made by site investigators who are movement disorder specialists. Data used in preparation for this article were obtained on February 13^th^, 2024 from the PPMI database (http://www.ppmi-info.org/access-data-specimens/download-data), RRID:SCR_006431. For up- to-data information on the study, visit http://www.ppmi-info.org.

External validation data came from the Systemic Synuclein Sampling Study (S4) cohort. S4 was a cross-sectional study evaluating a-syn measurement in tissue and biofluid samples among people with and without PD. Enrollment occurred between October 2015 and August 2017 at six sites in North America. The PD group included those with a decreased striatal dopamine transporter binding on SPECT (based on age-matched normative data), while the non-PD (‘healthy control’) group had normal DAT SPECT. Additional details about the S4 study design have been described elsewhere.^17,18^

### a-Syn Seed Amplification Assay (SAA)

a-Syn detection in the PPMI study was measured in CSF samples using the Amprion a-syn SAA, details of which have been previously described.^15,19^ Initial a-syn SAA testing in PPMI occurred in 2022 using an assay with a runtime length of 150 hours to produce aggregation curves for kinetic analysis of a-syn (PPMI biospecimen analysis project 155). In 2023, a-syn testing in PPMI was conducted using a more optimized assay that had a reduced runtime length of 24 hours and a reduced required volume of recombinant a-syn (PPMI biospecimen analysis project 237). For both assay versions, sample reactions were run in triplicate, and a final qualitative determination of being a-syn SAA positive, negative, or inconclusive was determined using maximum fluorescence (F_max_) data from all three replicates as described previously.^15,19^ Briefly, a sample was considered a-syn positive when all three replicates were positive according to a probabilistic algorithm based on F_max_. A sample was considered a-syn negative when zero or one replicates were positive, and was inconclusive when two replicates were positive. There were 1384 out of 3874 PPMI participants with CSF a-syn SAA data at the time of download. Data were excluded from the analysis for participants who had inconclusive results, or with re-tested samples showing inconsistent a-syn SAA results.

In the S4 study, cyclic amplification of protein misfolding adapted for a-syn was performed using CSF samples as previously described.^20^ Samples were run in triplicate. For each participant, the average F_max_ value from three samples was calculated to classify a-syn SAA status as positive or negative. Receiver-operating curve analysis identified an optimal F_max_ cutoff of ≥ 1375 for SAA status, based on the maximum Youden Index for differentiating PD from healthy controls.^21^ There were 75 out of 82 S4 participants with CSF a-syn SAA data.

### Clinical Demographics and Assessments Included in Analyses

To predict a-syn SAA positive versus negative status, we selected clinical and genetic variables that demonstrated subgroup differences in SAA status according to prior analysis.^15^ These variables included presence or absence of *LRRK2* pathogenic variants (regardless of PD manifestation) and the *LRRK2* variant type located within the kinase domain (G2019S or I2020T) versus the Ras of complex (ROC) proteins domain (R1441G/C or N1437H); any *GBA* pathogenic variant; University of Pennsylvania Smell Identification Test (UPSIT) scores; and, age and sex. We also evaluated constipation symptoms as a variable, which is a symptom that has been suggested to at least partially stem from a-syn accumulation in myenteric neurons.^22,23^ Constipation was measured by the Scale for Outcomes in PD – Autonomic Dysfunction constipation symptom (SCOPA-AUT5) categorized as never/sometimes or regular/often.

UPSIT data were utilized in multiple formats with varying criteria for missingness among the forty different scent items to assess the impact on modeling performance. First, we evaluated UPSIT data as age- and sex-specific percentiles, as described previously.^24^ This approach included participants missing at most one of the forty scent items. Missing responses were imputed as one, assuming correct identification of the scent; analyses were repeated with an imputation as zero, assuming an incorrect response. Results using an imputation of zero were nearly identical to those using an imputation of one, and therefore are not reported. We also used UPSIT data in a more general approach by calculating the percentage of correctly answered scent items without age and sex-specific scores. This UPSIT percent correct method included participants who completed more than twenty of the forty individual scent test items, and was calculated by dividing the total number of correct responses by the total number of items answered. Results for the UPSIT percent correct approach are provided in the Appendix.

### Statistical Analysis

Logistic regression models were developed using PPMI data that included participants from all cohorts (as described above) to predict the probability of SAA a-syn positive status. Candidate models were evaluated as follows: a null model without covariates; a baseline model that uses only UPSIT data; partially adjusted models that include UPSIT data plus age and/or sex; another partially adjusted model including UPSIT data, sex (or age), and constipation history (SCOPA-Aut item 5); and, a fully adjusted model that includes UPSIT data, sex (or age), constipation history, *LRRK2* pathogenic variant type, and *GBA* status. The relative goodness of fit between the nested models was assessed using Likelihood Ratio Chi-Squared tests. A significance level of p < 0.05 was used to determine whether the inclusion of additional predictors significantly improved the model fit. Regression coefficients from each logistic model and their odds ratios are reported to indicate the relative risk of SAA positive status associated with each predictor.

The predictive performance of the models was first examined through internal validation using area under the Receiver Operating Characteristic curve (AUROC). The optimal probability cut-off for defining SAA status for each model was determined using the Youden Index, maximizing the sum of sensitivity and specificity, from the internally trained models on PPMI data. Confusion matrices corresponding to the cut-offs were generated, and metrics such as sensitivity, specificity, accuracy, positive predictive value, and negative predictive value were calculated.

For internal validation, an optimism bootstrap validation approach on the PPMI dataset was implemented. We resampled, refit, and reevaluated on 10000 bootstrap re-samples. For external validation, the logistic regression model trained on the full PPMI dataset was subsequently applied to the independent S4 data cohort. Performance metrics in the external S4 sample were assessed to evaluate how well the models built on the PPMI cohort generalized to an external cohort that also consisted of participants with PD and healthy controls.

The models were also evaluated for their ability to predict CSF a-syn SAA status when considering only participants with manifest PD to determine their capabilities in a PD-specific setting. For these analyses, model performance metrics were re-calculated as previously described but when considering only those individuals who had a PD diagnosis at enrollment in either PPMI or S4.

Descriptive statistics were calculated for PPMI and S4 cohorts. Categorical variables were compared using chi-squared test of independence, and numeric variables were compared using Welch’s two-sample t-tests. Analyses were conducted in R 4.3.1 (packages tidyverse, pROC, and rms).

## RESULTS

### Model Development

Among the 1384 participants in the PPMI study who had CSF a-syn SAA status assayed, 1260 (91.0%) and 1313 (94.9%) participants met the data inclusion criteria when considering age- and sex-specific UPSIT percentile, and UPSIT percent correct, respectively (**Appendix Figure 1**). Forty-three participants were excluded due to inconsistent a-syn SAA status results, and four were excluded because of inconclusive results. An additional 23 individuals were excluded for missing > 50% of their UPSIT items, and one person did not have SCOPA-AUT5 data. Fifty-three participants who were missing more than one UPSIT item were excluded from the age- and sex-specific UPSIT percentile analysis, while 143 participants required imputation of one UPSIT item for this analysis. The baseline characteristics of participants included in the age- and sex-specific UPSIT percentile modeling analysis are summarized in **Table 1** (see **Appendix Table 1** for baseline characteristics from UPSIT percent correct analysis).

**Table 1.**
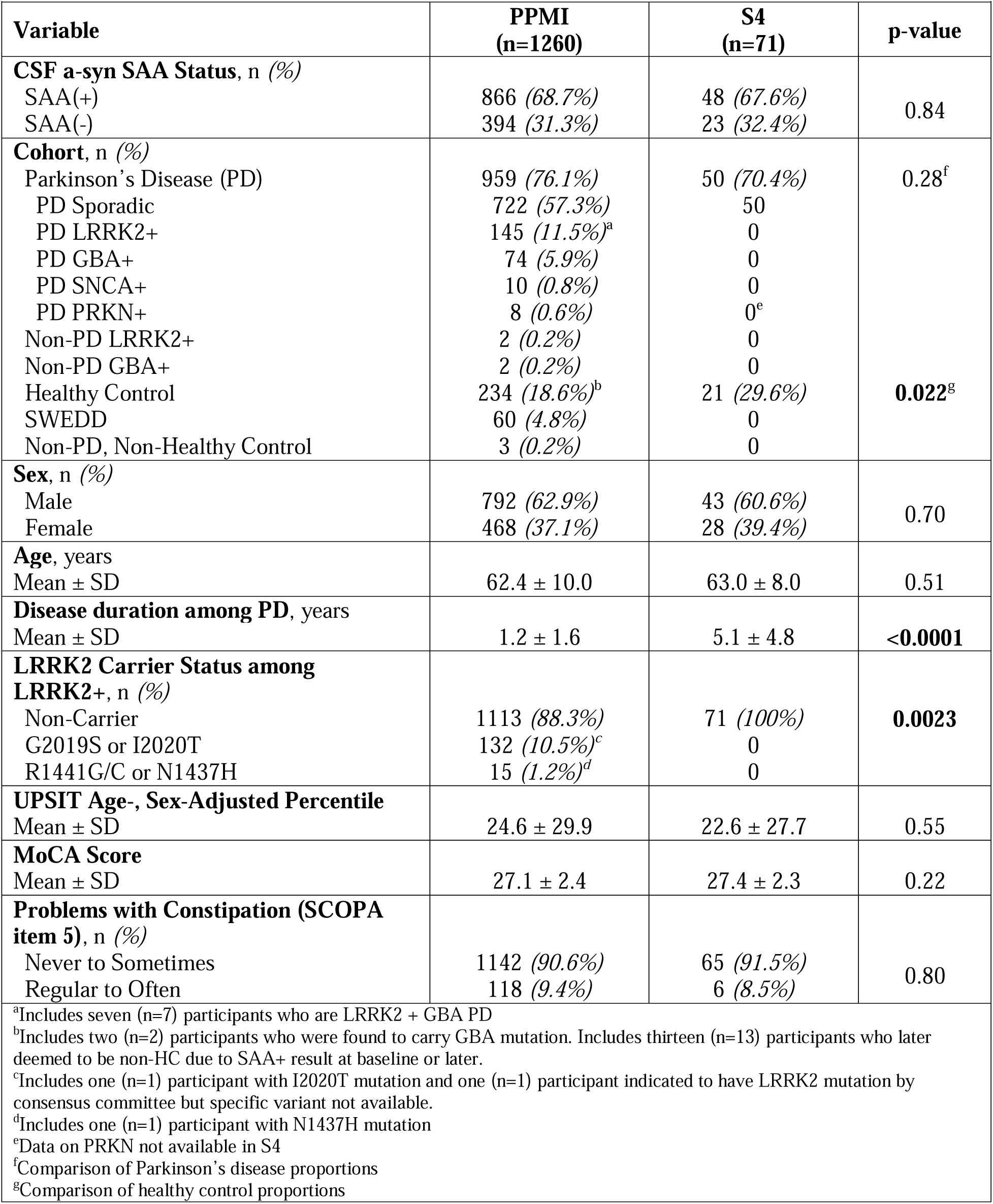
Demographics and clinical characteristics of PPMI (training) and S4 (validation) datasets.

For the candidate logistic regression models that used age- and sex-specific UPSIT percentiles, incorporation of sex, constipation symptoms, and *LRRK2* and *GBA* genetic information significantly improved model fit, as evidenced by the likelihood ratio tests (**Appendix Table 2**). For the logistic regression models that used UPSIT percent correct, the inclusion of age but not sex significantly improved model fit, while inclusion of constipation symptoms and genetics data remained beneficial (**Appendix Table 3**).

**Table 2.**
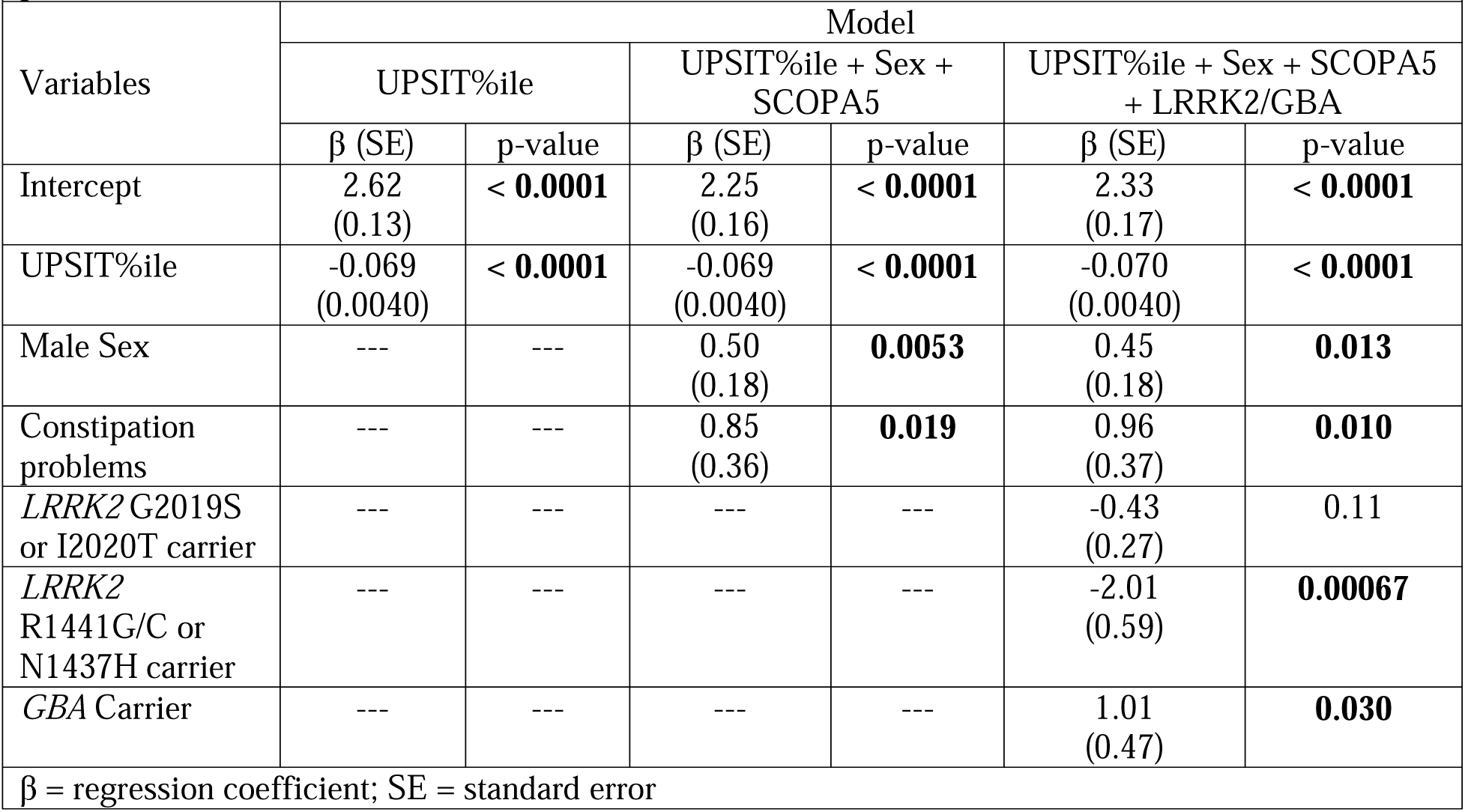
Logistic regression coefficient estimates from models using age- and sex-specific UPSIT percentile.

**Table 3.**
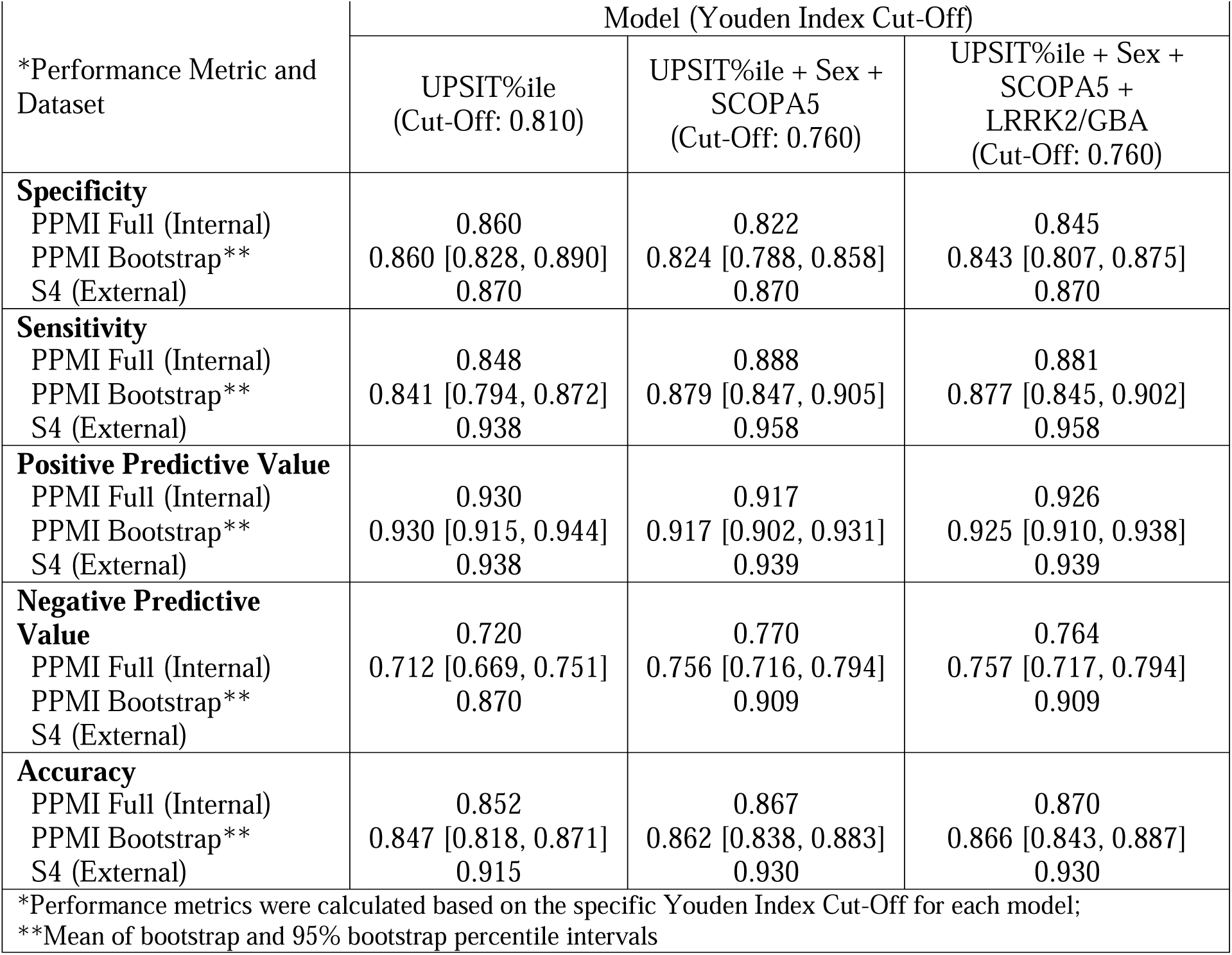
Internal and external model performances to discriminate a-Syn SAA+ vs. SAA-status using age- and sex-specific UPSIT percentile.

| *Performance Metric and Dataset | Model (Youden Index Cut-Off) |  |  |
| --- | --- | --- | --- |
|  | UPSIT%ile<br>(Cut-Off: 0.810) | UPSIT%ile + Sex + SCOPA5<br>(Cut-Off: 0.760) | UPSIT%ile + Sex + SCOPA5 + LRRK2/GBA<br>(Cut-Off: 0.760) |
| **Specificity** |  |  |  |
| PPMI Full (Internal) | 0.860 | 0.822 | 0.845 |
| PPMI Bootstrap** | 0.860 [0.828, 0.890] | 0.824 [0.788, 0.858] | 0.843 [0.807, 0.875] |
| S4 (External) | 0.870 | 0.870 | 0.870 |
| **Sensitivity** |  |  |  |
| PPMI Full (Internal) | 0.848 | 0.888 | 0.881 |
| PPMI Bootstrap** | 0.841 [0.794, 0.872] | 0.879 [0.847, 0.905] | 0.877 [0.845, 0.902] |
| S4 (External) | 0.938 | 0.958 | 0.958 |
| **Positive Predictive Value** |  |  |  |
| PPMI Full (Internal) | 0.930 | 0.917 | 0.926 |
| PPMI Bootstrap** | 0.930 [0.915, 0.944] | 0.917 [0.902, 0.931] | 0.925 [0.910, 0.938] |
| S4 (External) | 0.938 | 0.939 | 0.939 |
| **Negative Predictive Value** |  |  |  |
| PPMI Full (Internal) | 0.720 | 0.770 | 0.764 |
| PPMI Bootstrap** | 0.712 [0.669, 0.751] | 0.756 [0.716, 0.794] | 0.757 [0.717, 0.794] |
| S4 (External) | 0.870 | 0.909 | 0.909 |
| **Accuracy** |  |  |  |
| PPMI Full (Internal) | 0.852 | 0.867 | 0.870 |
| PPMI Bootstrap** | 0.847 [0.818, 0.871] | 0.862 [0.838, 0.883] | 0.866 [0.843, 0.887] |
| S4 (External) | 0.915 | 0.930 | 0.930 |
| *Performance metrics were calculated based on the specific Youden Index Cut-Off for each model; |  |  |  |
| **Mean of bootstrap and 95% bootstrap percentile intervals |  |  |  |

The parameter estimates from partially and fully adjusted models using age- and sex-specific UPSIT percentiles are reported in **Table 2**. Overall, the fully adjusted model indicates that higher odds of a-syn SAA(+) status are associated with a lower UPSIT age- and sex-specific percentile, male sex, the presence of constipation problems, and carrying a *GBA* mutation. In contrast, having a *LRRK2* variant, particularly R1441G/C or N1437H, is associated with lower odds of a-syn SAA(+) status. For the fully adjusted model using UPSIT percent correct, age, constipation history, and *LRRK2* and *GBA* genetic information, the general patterns were similar (**Appendix Table 4**); however, the coefficient estimate (β) for the UPSIT percent correct was larger in magnitude (β = -0.12), and increasing age was associated with lower odds of a-syn SAA(+) status (β = -0.047).

The odds ratios (ORs) of the fully adjusted logistic regression model using age- and sex-specific UPSIT percentiles are presented in **Figure 1**. The a-syn SAA(+) OR for UPSIT age- and sex-specific percentile is 0.932, indicating that between two individuals whose only difference is a 1-percentile difference in UPSIT, the individual in the higher percentile has 0.932 times the odds of being a-syn SAA(+). Rescaling this OR for 10-percentile unit changes shows that for someone whose age- and sex-specific UPSIT is higher by 10 percentile, they will have 0.497 times the odds of being a-syn SAA(+). Carrying the R1441G/C *LRRK2* variant was another significant impactful variable, with carriers having 0.134 times the odds of being a-syn SAA(+) compared to non-carriers. In contrast, *GBA* mutation carriers had 2.8 times the odds of being a-syn SAA(+) compared to non-*GBA* mutation carriers. Male sex compared to female sex, and more frequent constipation problems versus no or less constipation problems were associated with 1.6 and 2.6 higher odds, respectively, for being a-syn SAA(+). A similar OR plot is provided in the Appendix for the UPSIT percent correct fully adjusted model (**Appendix Figure 2**).

**Figure 1.**
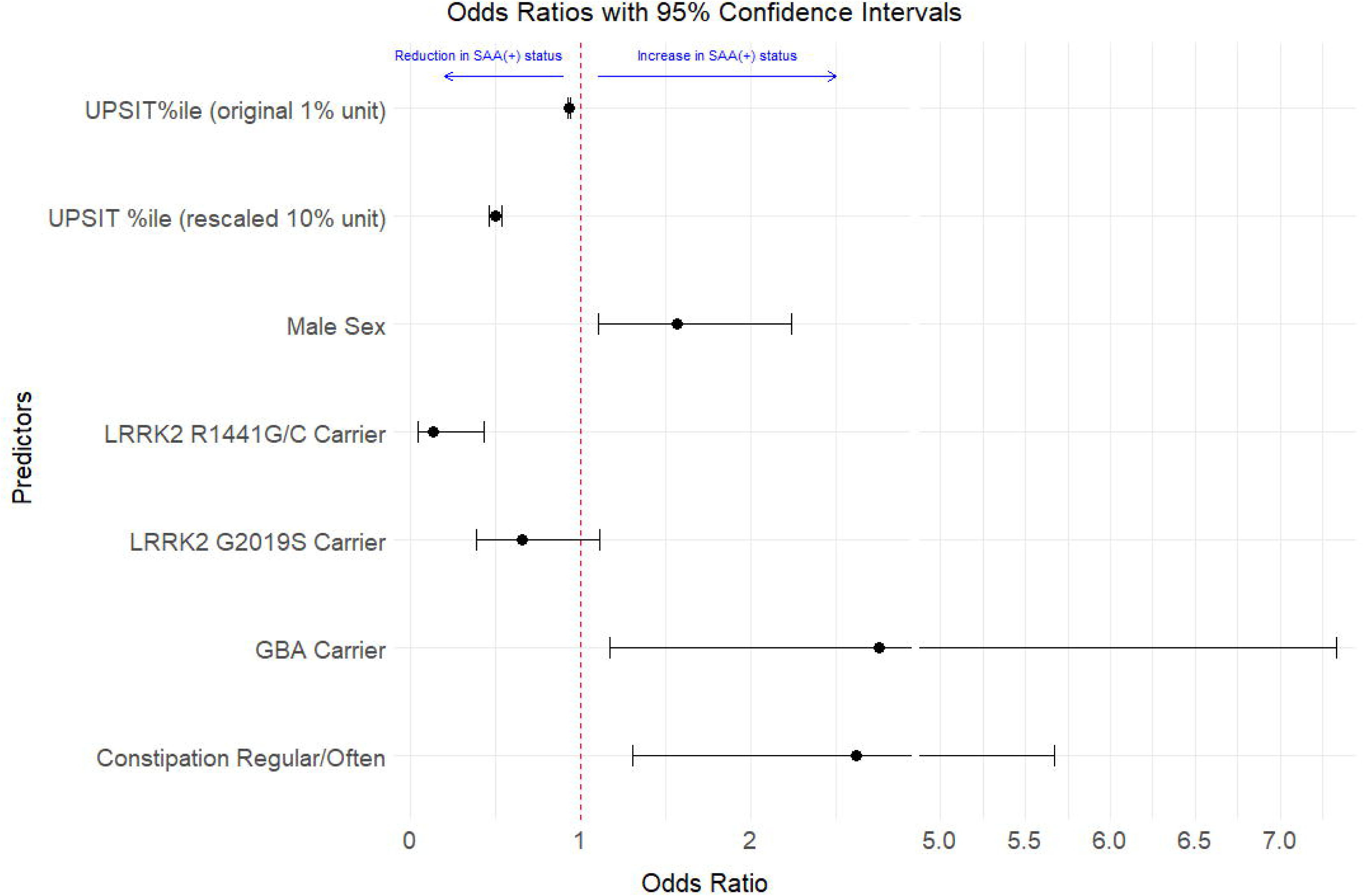
Odds Ratios (with 95% Confidence Intervals) from Fully Adjusted Logistic Regression Model. Values less than one are associated with a reduced risk of positive alpha-synuclein results from the cerebrospinal fluid seeding amplification assay (a-syn SAA+), and values greater than one are associated with an increased risk of a-syn SAA+, relative to the average population sample. UPSIT percent correct are plotted using 1% and 10% unit changes. Plotted with ‘ggbreak’ R package.^25^

Alternatively, log-odds can be viewed as probabilities for additional interpretations. In **Figure 2**, we provide probability plots for a-syn SAA(+) status versus UPSIT age- and sex-specific percentiles by constipation status and genetic status among males (**Figure 2A**) and females (**Figure 2B**). To facilitate further exploration of the models, an interactive calculator is available online at https://chet-analytics.shinyapps.io/saa_r_shiny/, allowing users to calculate the probabilities of a-syn SAA(+) by entering values of the predictor variables.

**Figure 2.**
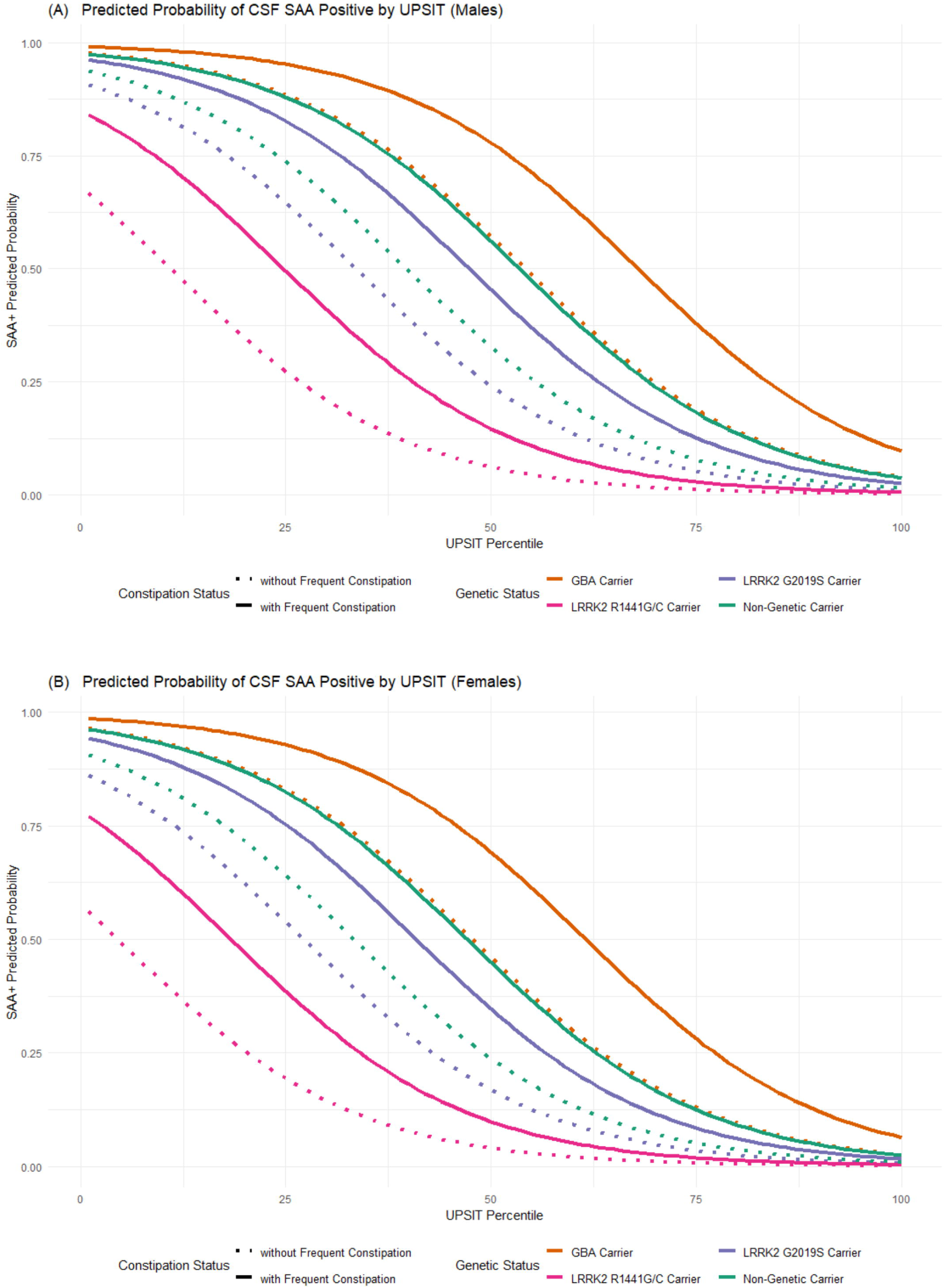
Probability of a-syn SAA(+) versus UPSIT age- and sex-specific percentile according to the fully adjusted model for (A) males and (B) females. Each line is shown for individuals who have a specific LRRK2 pathogenic variant, or carry GBA pathogenic variant, or do not carry those specific LRRK2 or GBA variants, while varying constipation status.

### Model-Based Predictions: Internal Performance

Using UPSIT age- and sex-specific percentiles to predict a-syn SAA status within the PPMI cohort, both the partially- and fully-adjusted models demonstrated high discrimination abilities in internal testing, as shown by their ROC curves (**Figure 3A**). Similar performance was observed when using UPSIT percent correct (**Appendix Figure 3**).

**Figure 3.**
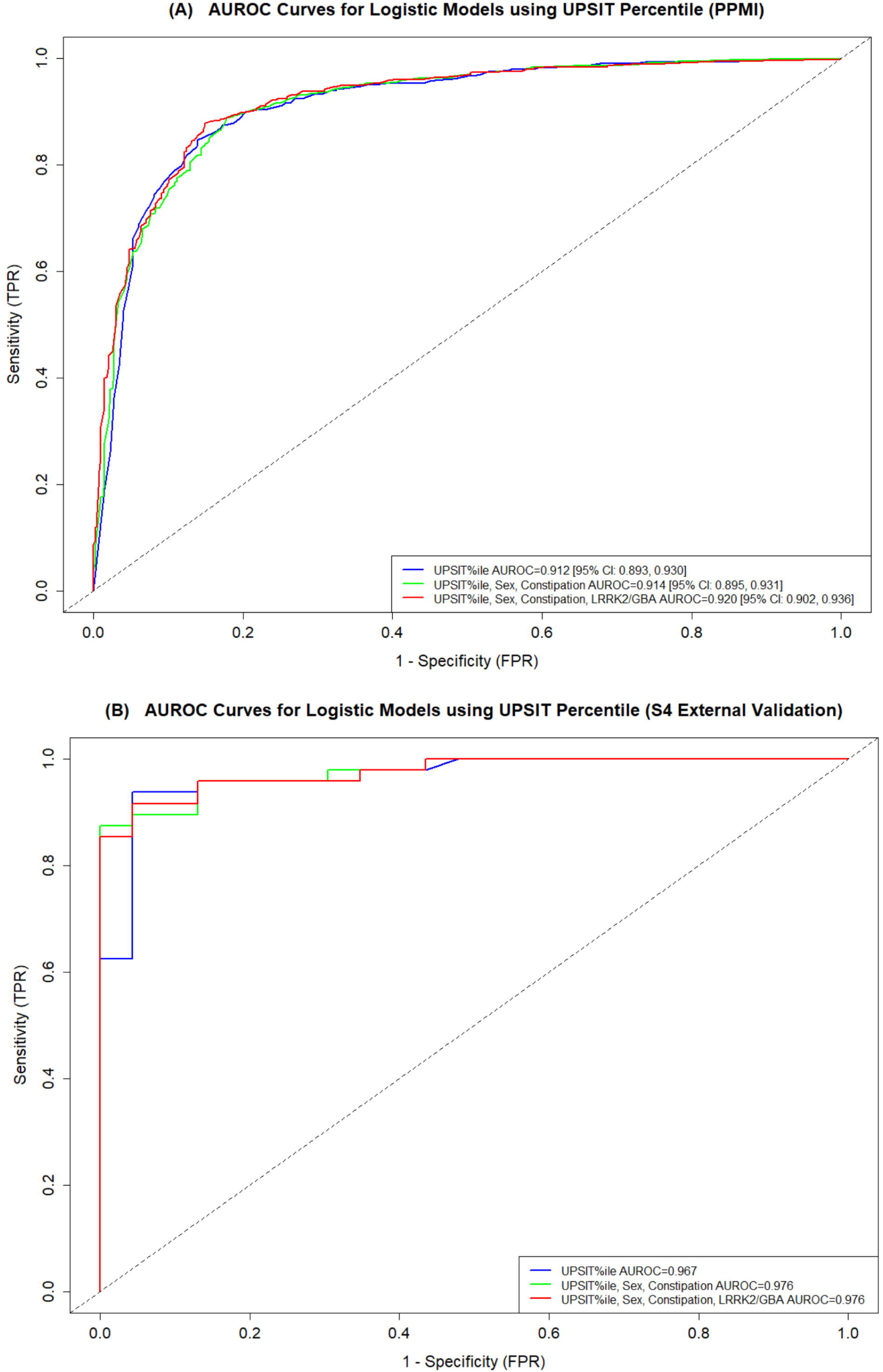
Training and validation sample AUROC curves from partially and fully adjusted models using UPSIT age- and sex-specific percentile to predict a-syn SAA status. The figures show the (A) PPMI training sample AUROC curves and values with 95% Confidence Intervals [95% CI] from the bootstrap analysis, and (B) the S4 validation sample AUROC curves.

Applying cut-offs optimized for sensitivity and specificity, the probabilities derived from the models were assessed for accuracy, sensitivity, specificity, positive predictive value, and negative predictive value in determining a-syn SAA(+) versus SAA(-) status (**Table 3**). Results from the internal and optimized bootstrap PPMI data demonstrate that all of the models performed well in predicting a-syn SAA status, with the fully adjusted model achieving the highest accuracy. Results were similar when using UPSIT percent correct instead of UPSIT age- and sex-specific percentiles (**Appendix Table 5**).

### Model-Based Predictions: External Performance

For external validation, the models constructed from the PPMI data were used to predict a-syn SAA status in the S4 cohort. The S4 dataset included 75 participants with a-syn SAA data; however, 71 participants were ultimately included because 4 were also co-enrolled in the PPMI study and were therefore excluded to prevent data leakage between the model training/testing and external validation datasets. Differences between the PPMI and S4 cohorts included a longer disease duration for PD participants in the S4 dataset, a higher percentage of healthy controls in S4, and no identified *LRRK2* or *GBA* pathogenic carriers in S4 (**Table 1**).

The partially- and fully-adjusted models trained on PPMI data maintained strong discrimination abilities in predicting a-syn SAA status in the S4 cohort when using UPSIT age- and sex-specific percentiles (**Figure 3B**) or UPSIT percent correct (**Appendix Figure 3**). Applying the same Youden Index cut-offs from the PPMI models to predict a-syn SAA(+) versus SAA(-) status in S4, the partially- and fully-adjusted models demonstrated strong performances in distinguishing between the groups (**Table 3; Appendix Table 5**).

### Model-Based Predictions: PD-Only

Using the partially- and fully-adjusted models, performance metrics were recalculated for the PPMI and S4 datasets, focusing solely on participants with manifest PD at enrollment, in contrast to a group comprised of both with, without, and at risk for PD. In the PPMI dataset, this comprised 966 participants, of whom 87.5% were a-syn SAA(+). In the S4 dataset, the subset included 50 participants, with 96.0% being a-syn SAA(+). **Table 4** shows the performance metrics of the three different models in distinguishing a-syn SAA(+) from SAA(-) status using age- and sex-specific UPSIT percentile-based models. The consistently high sensitivities, PPVs, and accuracies across datasets for all models indicate they remained effective in detecting positive cases. However, there were decreases in specificity and NPV indicating a diminished ability to rule out negative cases among manifest PD participants. Results were similar when using UPSIT percent correct (**Appendix Table 6**).

**Table 4.** Model performance metrics among manifest PD participants to discriminate a-Syn SAA+ vs. SAA-status using age- and sex-specific UPSIT percentile.

| *Performance Metric and Dataset | Model (Youden Index Cut-Off) |  |  |
| --- | --- | --- | --- |
|  | UPSIT%ile<br>(Cut-Off: 0.810) | UPSIT%ile + Sex +<br>SCOPA5<br>(Cut-Off: 0.760) | UPSIT%ile + Sex +<br>SCOPA5 +<br>LRRK2/GBA<br>(Cut-Off: 0.760) |
| **Specificity** |  |  |  |
| PPMI (Internal) | 0.785 | 0.711 | 0.785 |
| S4 (External) | 0.500 | 0.500 | 0.500 |
| **Sensitivity** |  |  |  |
| PPMI (Internal) | 0.853 | 0.893 | 0.885 |
| S4 (External) | 0.935 | 0.957 | 0.957 |
| **Positive Predictive Value** |  |  |  |
| PPMI (Internal) | 0.965 | 0.956 | 0.966 |
| S4 (External) | 0.956 | 0.957 | 0.957 |
| **Negative Predictive Value** |  |  |  |
| PPMI (Internal) | 0.434 | 0.489 | 0.495 |
| S4 (External) | 0.400 | 0.500 | 0.500 |
| **Accuracy** |  |  |  |
| PPMI (Internal) | 0.845 | 0.871 | 0.873 |
| S4 (External) | 0.900 | 0.920 | 0.920 |
| *Performance metrics were calculated based on the specific Youden Index Cut-Off for each model |  |  |  |

## DISCUSSION

This study explores the potential of data-driven models to predict CSF a-syn SAA status in a group of individuals either with or at-risk for PD using readily available clinical data. We found that models using UPSIT data, sex, constipation history, and simple genetics information can predict CSF a-syn SAA status with good accuracy. The predictive capability of the models, demonstrated through AUROC values exceeding 0.900 in both internal and external validation cohorts, highlights the robustness of the models in PD-enriched populations. Interestingly, models that only included UPSIT data showed only marginally reduced performance compared to the partially- and fully-adjusted models. These results underscore the feasibility of leveraging non-invasive measures to help infer the likelihood of the presence of seeding-capable a-syn as detected by CSF a-syn SAA.

Our models suggests that olfactory dysfunction is a useful and potent indicator of CSF a-syn SAA status, reinforcing its significance as an early marker in neurodegenerative processes, particularly in PD. Previous research demonstrate that olfactory dysfunction predicts conversion to PD and dementia with Lewy bodies among patients with isolated REM sleep behavior disorder (iRBD).^26-28^ For example, Coughlin and colleagues reported that age- and sex-specific UPSIT percentile had the highest predictive capacity among a range of clinical and demographic features, with an AUROC of 0.87 in predicting CSF a-syn positivity among participants with dementia with Lewy bodies.^29^ Furthermore, significant associations have been observed between a-syn pathology and worse olfactory function in PD, iRBD, and normal aging populations.^30-33^

The olfactory tract is considered an initiation site for the seeding and spread of pathological a-syn in PD. Recent disease progression modeling of 814 brain donors indicates that most with Lewy body pathology show earliest pathology in the olfactory bulb, while a smaller subset shows later olfactory involvement after initial brainstem pathology and spread.^34^ Notably, those inferred to have a spatiotemporal trajectory involving later olfactory bulb pathology but earlier brainstem pathology and more widespread regional brain involvement had worse pre-mortem smell dysfunction compared to those inferred to have early olfactory bulb pathology, suggesting this symptom is linked to overall brain pathology rather than localized changes. Clinicopathological studies in non-PD populations support the notion that olfactory dysfunction could be more associated with widespread Lewy body pathology rather than localized olfactory changes.^32,33^ However, there are experimental studies in mice that challenge this view, demonstrating that olfactory dysfunction can occur due to localized a-syn aggregation in the olfactory bulb by disrupting neural activity and synaptic transmission.^35^ Additionally, RNA sequencing has revealed significant gene alterations in the olfactory bulb of later-stage PD patients, implicating pathways related to neuroinflammation, neurotransmitter dysfunction, and disruptions in olfactory transduction signaling.^36^ While the exact mechanisms of olfactory dysfunction in PD will require further investigation, we clearly show that olfactory dysfunction is a strong indicator of CSF a-syn SAA positivity in PD.

Our modeling also found that *LRRK2* pathogenic carriers had reduced odds of being a-syn CSF SAA positive compared to PD non-carriers, while *GBA* pathogenic carriers had increased odds of positivity. In the PPMI training dataset, 63% of *LRRK2* PD pathogenic carriers had positive a-syn CSF SAA results, while 93% of the sporadic PD population were positive. This aligns with prior research reporting 40% to 78% of *LRRK2* carriers having positive a-syn seeding activity compared to approximately 90% of sporadic PD participants.^15,37,38^

Our models suggest that the type of *LRRK2* pathogenic variant is also important to consider when predicting a-syn SAA status. Carriers of R1441G/C/N1437H had the greatest reduction in odds of being a-syn CSF SAA positive compared to non-carriers, while G2019S/I2020T carriers also had reduced odds but to a lesser extent. Several neuropathological studies of R1441G/C/H PD carriers have shown that Lewy body pathology is absent or rarely present at autopsy.^39-41^ Histopathology from individuals carrying the G2019S mutation seem to show a-syn pathology is more common but is still highly variable, which aligns with our modeling.^42,43^ For instance, among 37 *LRRK2*-related PD cases, all reporting neuronal loss within the substantia nigra, Lewy body pathology was present in 11/17 (65%) G2019S mutation carriers, and 2/6 (33%) R1441C/G carriers.^44^ It is unclear as to the specific biochemical mechanism(s) that could be driving these differences in a-syn seeding activities between different *LRRK2* pathogenic variants, and it will be important to investigate these differences further for potential therapies that target a-syn aggregation and *LRRK2* carriers.

A history of constipation symptoms self-reported as “regular” or “often” was also a significant predictor of CSF a-syn SAA positive status in the fully adjusted model. Individuals with more frequent constipation had more than 2.5 times higher odds of being a-syn SAA(+) than those with no or infrequent constipation issues. Constipation, a common non-motor symptom of PD, is also a predictor of developing the disease.^45^ Neuropathological studies have shown a-syn accumulation in the enteric nervous system in both PD and Lewy body disease, and some experimental studies suggest that constipation may be related to the formation of a-syn in the GI tract.^23,46^ However, few clinical studies have directly measured the association of a-syn pathology with constipation. While immunohistochemistry studies have shown minimal evidence linking localized Lewy pathology in colonic and mucosal biopsies with constipation in PD and non-PD subjects, a-syn seeding assays have demonstrated an association.^22^ Kuzkina and colleagues found higher a-syn seeding activity from skin biopsy samples in PD patients with constipation compared to those without.^8^ Thus, incorporating a simple self-reported history of constipation symptoms questionnaire seems to be a plausible and helpful predictor in identifying some individuals at-risk of having positive a-syn SAA results.

Male sex was associated with higher odds of a-syn SAA(+) results when using age- and sex-specific UPSIT percentiles, but it was not a significant predictor in models that used UPSIT percent correct scores. Conversely, increasing age was linked to lower odds of being a-syn SAA(+) when using UPSIT percent correct scores, but this association was not observed with the age- and sex-adjusted percentiles. These findings suggest that the influence of sex and age on a-syn status may be driven more by the method in which UPSIT data are incorporated into the models rather than by true biological differences. Nevertheless, it would be interesting to investigate further how these variables interact and whether different approaches to incorporating UPSIT data could reveal more nuanced insights into the relationship between sex, age, and a-syn pathology.

These models could be useful in several ways. First, they could facilitate identification of individuals who would most benefit from CSF a-syn SAA testing in the research setting, optimizing resource allocation, and minimizing unnecessary invasive procedures. For example, if an individual has a very low probability of being a-syn SAA(+) according to the models, they may not need further invasive testing. Additionally, these models can be applied to historical datasets that lack CSF samples but have UPSIT data, allowing researchers to understand the influence of a-syn SAA status on various exploratory analyses. Furthermore, the models can be utilized in research settings where CSF sample collection is not feasible. This flexibility enables researchers to infer SAA status likelihood across a broader spectrum of PD studies, potentially enriching our understanding of disease patterns and treatment responses in more diverse patient populations. By enhancing the ability to estimate a-syn pathology without direct sample analysis, these models promise to expand the scope and applicability of this area of PD research.

Our study has some limitations. The external validation cohort was relatively small compared to the training dataset, and did not include participants with *LRRK2* or *GBA* PD-associated genetic mutations. Therefore, we were unable to truly test how well our fully-adjusted model could perform in the setting of a more genetically diverse PD population whose a-syn pathology is more variable. It would be of great interest to test and recalibrate these models using additional datasets that have collected CSF a-syn SAA data with UPSIT information in populations with PD and other neurodegenerative diseases. We also did not include rapid eye movement behavior disorder (RBD) as a potential predictor in our models, which has been shown to be a risk-factor for the presence a-syn pathology.^29^ While the PPMI study collected RBD data through the RBD Questionnaire, the S4 validation dataset did not. Therefore, we elected not to pursue the use of RBD symptoms in our models. There were also limitations of the models when using them specifically in a PD manifest population, particularly around the specificity and NPV of the models, indicating a higher rate of false positives. This suggests that while the models are effective at identifying true positives, they may still require further refinement to reduce false positive rates and improve their overall diagnostic utility when only considering manifest PD.

## DATA AVAILABILITY

The data supporting the findings from this study are available at the Parkinson’s Progression Marker Initiative (PPMI) website (https://www.ppmi-info.org/). This analysis used data openly available from the PPMI (Tier 1 data). Protocol information for The Parkinson’s Progression Markers Initiative (PPMI) Clinical - Establishing a Deeply Phenotyped PD Cohort AM 3.2. can be found on protocols.io or by following this link: https://dx.doi.org/10.17504/protocols.io.n92ldmw6ol5b/v2. Additional data from the Systemic Synuclein Sampling Study (S4) were available through Zenodo (https://zenodo.org/records/7821869) after registration for approved data access. Statistical analysis codes used to perform the analyses in this article are shared on Zenodo (https://doi.org/10.5281/zenodo.14867028).

