## Appendix for "Predicting Cerebrospinal Fluid Alpha-Synuclein Seed Amplification Assay Status from Demographics and Clinical Data"

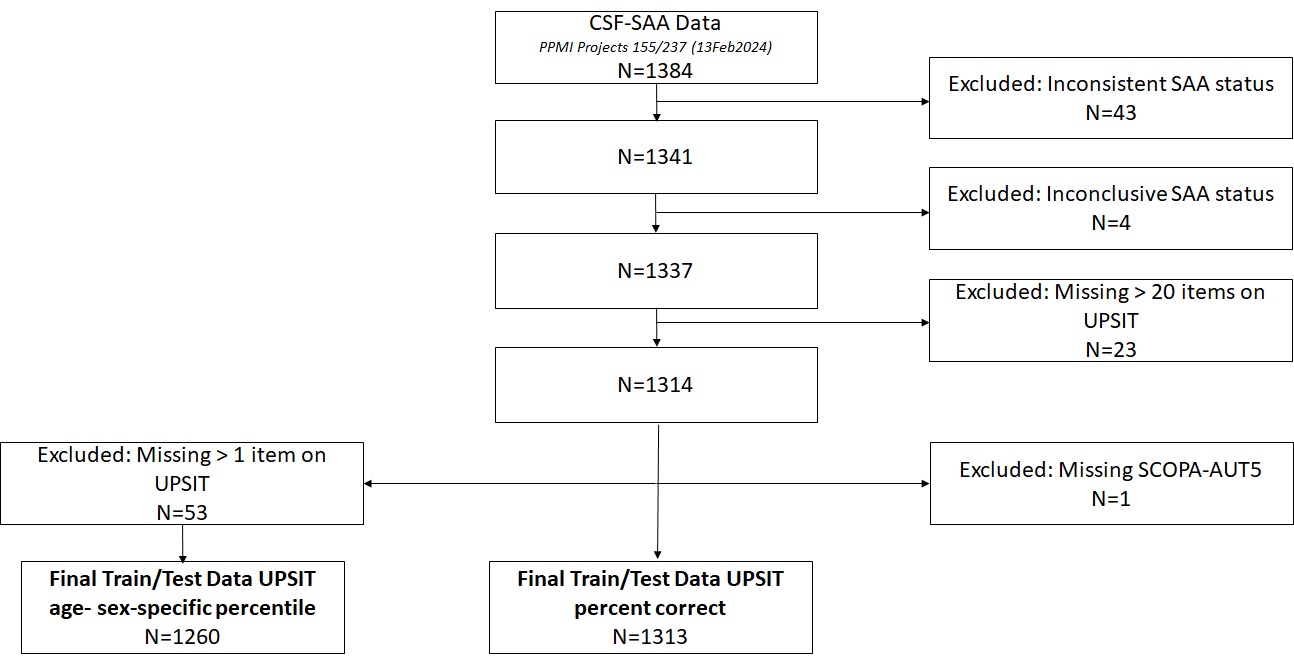

### Appendix Figure 1. Participant data included from PPMI in the model training/testing dataset.

| **Appendix Table 1. Demographics and clinical characteristics of PPMI (training) and S4 (validation) datasets when considering individuals > 20 UPSIT items answered** | | | |
| --- | --- | --- | --- |
| **Variable** | **PPMI**  **(n=1313)** | **S4**  **(n=71)** | **p-value** |
| **CSF a-syn SAA Status**, n *(%)*  SAA(+)  SAA(-) | 913 *(69.5%)*  400 *(30.5%)* | 48 *(67.6%)*  23 *(32.4%)* | 0.73 |
| **Cohort**, n *(%)*  Parkinson’s Disease (PD)  PD Sporadic  PD LRRK2+  PD GBA+  PD SNCA+  PD PRKN+  Non-PD LRRK2+  Non-PD GBA+  Healthy Control  SWEDD  Non-PD, Non-Healthy Control | 1005 *(76.5%)*  766 *(58.3%)*  145 *(11.0%)*^a^  74 *(5.6%)*  11 *(0.8%)*  9 *(0.7%)*  2 *(0.2%)*  2 *(0.2%)*  241 *(18.4%)*^b^  60 *(4.6%)*  3 *(0.2%)* | 50 *(70.4%)*  0  0  0  0  0^e^  0  0  21 *(29.6%)*  0  0 | 0.24^f^  **0.019**^g^ |
| **Sex**, n *(%)*  Male  Female | 822 *(62.6%)*  491 *(37.4%)* | 43 *(60.6%)*  28 *(39.4%)* | 0.73 |
| **Age**, years  Mean ± SD | 62.5 ± 10.0 | 63.0 ± 8.0 | 0.62 |
| **Disease duration among PD**, years  Mean ± SD | 1.2 ± 1.6 | 5.1 ± 4.8 | **<0.0001** |
| **LRRK2 Carrier Status among LRRK2+**, n *(%)*  Non-Carrier  G2019S or I2020T  R1441G/C or N1437H | 1166 *(88.8%)*  132 *(10.1%)^c^*  15 *(1.1%)^d^* | 71 *(100%)*  0  0 | 0.0029 |
| **UPSIT Percent Correct**  Mean ± SD | 62.4 ± 21.9 | 59.7 ± 22.2 | 0.32 |
| **MoCA Score**  Mean ± SD | 27.0 ± 2.5 | 27.4 ± 2.3 | 0.18 |
| **Problems with Constipation (SCOPA item 5)**, n *(%)*  Never to Sometimes  Regular to Often | 1191 *(90.7%)*  122 *(9.3%)* | 65 *(91.5%)*  6 *(8.5%)* | 0.81 |
| ^a^Includes six (n=6) participants who are LRRK2 + GBA PD  ^b^Includes two (n=2) participants who were found to carry GBA mutation. Includes fifteen (n=15) participants who later deemed to be non-HC due to SAA+ result at baseline or later.  ^c^Includes one (n=1) participant with I2020T mutation and one (n=1) participant indicated to have LRRK2 mutation by consensus committee but specific variant not available.  ^d^Includes one (n=1) participant with N1437H mutation  ^e^Data on PRKN not available in S4  ^f^Comparison of Parkinson’s disease proportions  ^g^Comparison of healthy control proportions | | | |

| **Appendix Table 2. Logistic regression likelihood ratio test of candidate using age- and sex-specific UPSIT percentile (imputing missing UPSIT items as one)** | | | |
| --- | --- | --- | --- |
| **Model #** | **Covariates** | **AIC** | **p-value** |
| 0 | Null (intercept only) | 1567.5 | -- |
| 1 | UPSIT%ile | 911.61 | **< 2e-16**  (vs. model 0) |
| 2 | UPSIT%ile + age | 913.41 | 0.66  (vs. model 1) |
| 3 | UPSIT%ile + sex | 906.82 | **0.0092**  (vs. model 1) |
| 4 | UPSIT%ile + sex + SCOPA5 | 902.48 | **0.012**  (vs. model 3) |
| 5 | UPSIT%ile + sex + SCOPA5 + LRRK2/GBA | 889.40 | **0.00026**  (vs. model 4) |

| **Appendix Table 3. Logistic regression likelihood ratio test of candidate models using UPSIT percent correct** | | | |
| --- | --- | --- | --- |
| **Model #** | **Covariates** | **AIC** | **p-value** |
| 0 | Null (intercept only) | 1616.3 | -- |
| 1 | UPSIT%ile | 951.03 | **< 2e-16**  (vs. model 0) |
| 2 | UPSIT%ile + age | 917.65 | **< 2.7e-09**  (vs. model 1) |
| 3 | UPSIT%ile + sex | 951.22 | 0.18  (vs. model 1) |
| 4 | UPSIT%ile + age + sex | 917.66 | 0.16  (vs. model 2) |
| 5 | UPSIT%ile + age + SCOPA5 | 913.21 | **0.011**  (vs. model 2) |
| 6 | UPSIT%ile + sex + SCOPA5 + LRRK2/GBA | 904.68 | **0.0023**  (vs. model 5) |

| **Appendix Table 4. Logistic regression coefficient estimates from models using UPSIT percent correct** | | | | | | |
| --- | --- | --- | --- | --- | --- | --- |
| Variables | Model | | | | | |
|  | UPSIT % Correct | | UPSIT % Correct + Age + SCOPA5 | | UPSIT % Correct + Age + SCOPA5 + LRRK2/GBA | |
|  | β (SE) | p-value | β (SE) | p-value | β (SE) | p-value |
| Intercept | 8.57  (0.47) | **< 0.0001** | 12.19  (0.84) | **< 0.0001** | 12.19  (0.85) | **< 0.0001** |
| UPSIT % Correct | -0.11 (0.0062) | **< 0.0001** | -0.12 (0.0068) | **< 0.0001** | -0.12  (0.0069) | **< 0.0001** |
| Age | --- | --- | -0.049  (0.0084) | **<0.0001** | -0.047  (0.0084) | **<0.0001** |
| Constipation problems | --- | --- | 0.85  (0.35) | **0.014** | 0.97  (0.36) | **0.0061** |
| LRRK2 G2019S or I2020T carrier | --- | --- | --- | --- | -0.47  (0.28) | 0.091 |
| LRRK2 R1441G/C or N1437H carrier | --- | --- | --- | --- | -2.01  (0.64) | **0.0016** |
| GBA Carrier | --- | --- | --- | --- | 0.60 (0.46) | 0.19 |
| β = regression coefficient; SE = standard error | | | | | | |

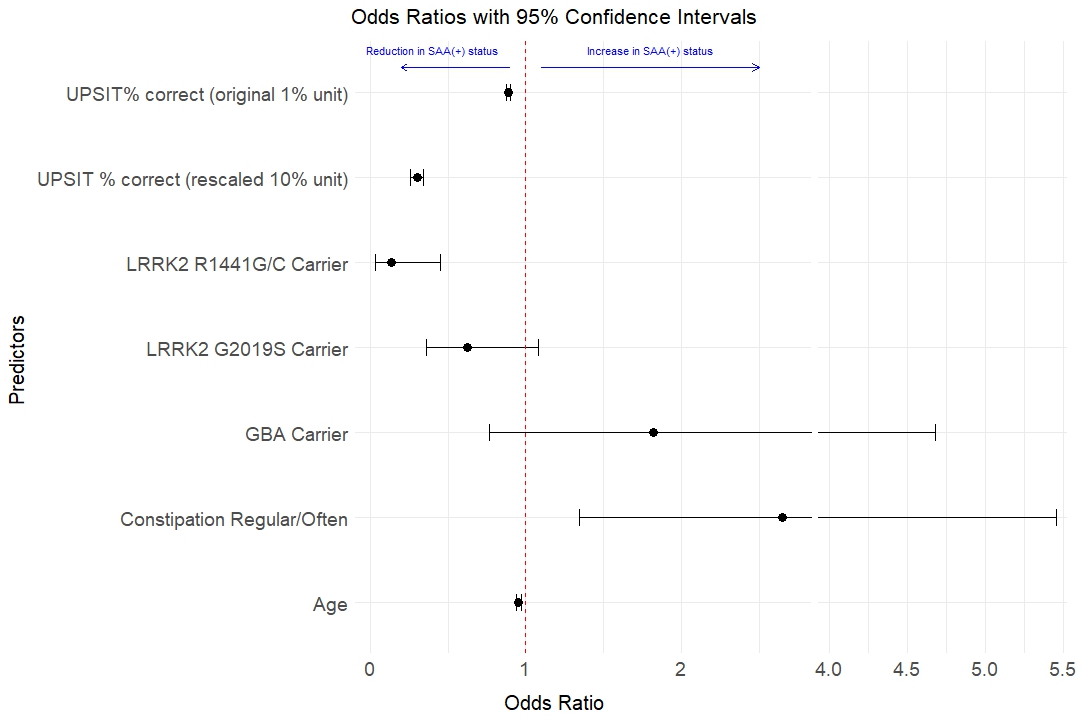

Appendix Figure 2. Odds Ratios (with 95% Confidence Intervals) from fully adjusted logistic regression model using UPSIT percent correct. Values less than one are associated with a reduced risk of positive alpha-synuclein results from the cerebrospinal fluid seeding amplification assay (a-syn SAA+), and values greater than one are associated with an increased risk of a-syn SAA+, relative to the average population sample. UPSIT percent correct are plotted using 1% and 10% unit changes.

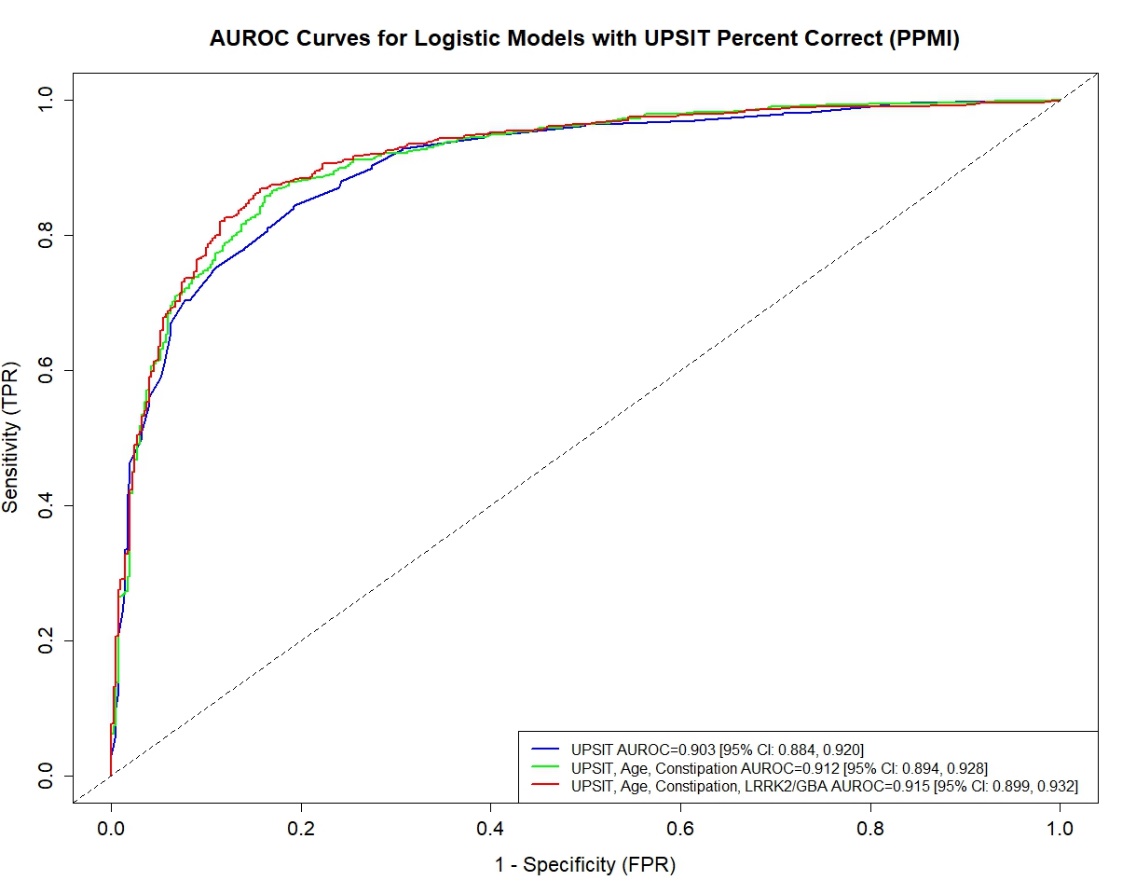

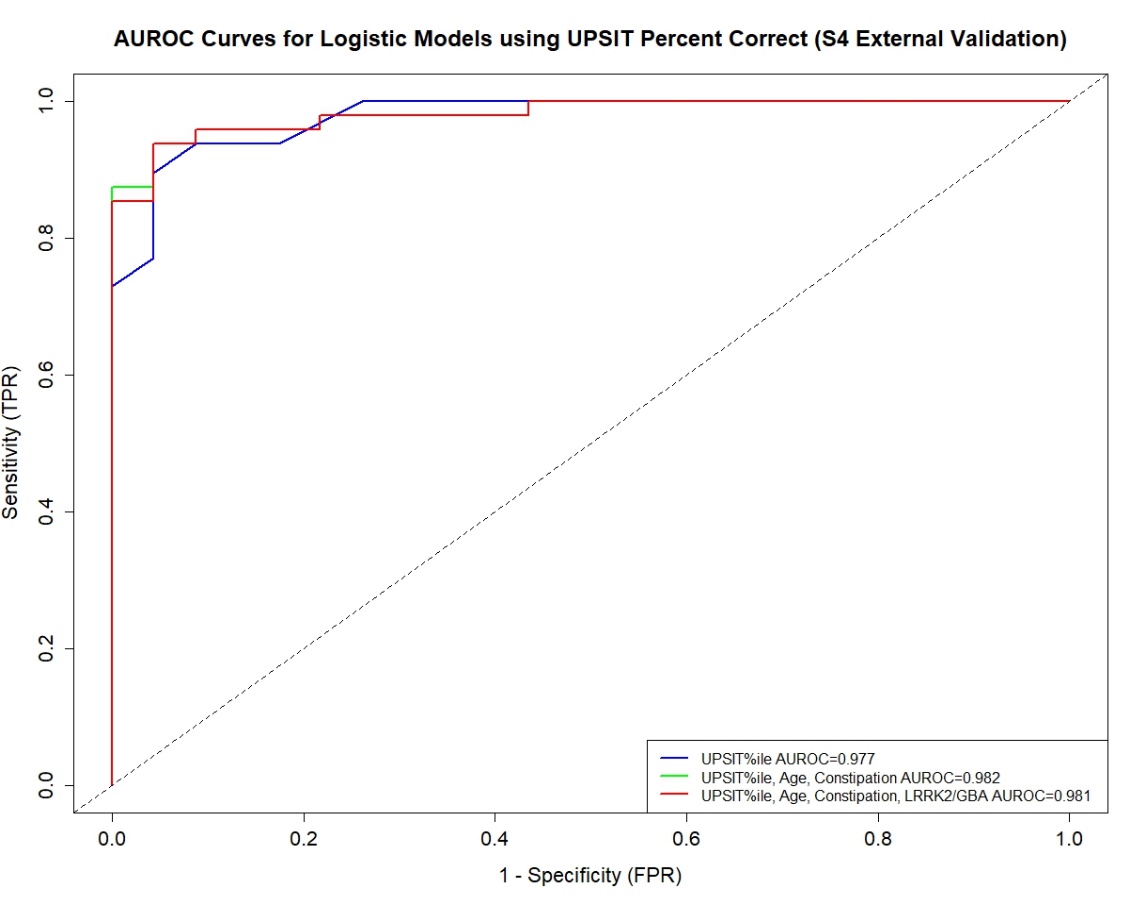

Appendix Figure 3. Training and validation sample AUROC curves from partially and fully adjusted models using UPSIT percent correct. The figures show the (Top) PPMI training sample AUROC curves and values with 95% Confidence Intervals [95% CI] from the bootstrap analysis, and (Bottom) the S4 validation sample AUROC curves.

| **Appendix Table 5. Internal and external model performances to discriminate a-Syn SAA+ vs. SAA- status with models using UPSIT percent correct** | | | |
| --- | --- | --- | --- |
| *Performance Metric and Dataset | Model (Youden Index Cut-Off) | | |
|  | UPSIT % Correct  (Cut-Off: 0.600) | UPSIT % Correct + Age + SCOPA5  (Cut-Off: 0.610) | UPSIT % Correct + Age + SCOPA5 + LRRK2/GBA  (Cut-Off: 0.600) |
| **Specificity**  PPMI Full (Internal)  PPMI Bootstrap**  S4 (External) | 0.808  0.799 [0.753, 0.838]  0.870 | 0.838  0.831 [0.795, 0.863]  0.913 | 0.843  0.839 [0.801, 0.873]  0.870 |
| **Sensitivity**  PPMI Full (Internal)  PPMI Bootstrap**  S4 (External) | 0.843  0.847 [0.823, 0.873]  0.938 | 0.858  0.859 [0.834, 0.882]  0.958 | 0.865  0.867 [0.843, 0.889]  0.958 |
| **Positive Predictive Value**  PPMI Full (Internal)  PPMI Bootstrap**  S4 (External) | 0.909  0.906 [0.889, 0.923]  0.938 | 0.923  0.921 [0.904, 0.935]  0.958 | 0.926  0.925 [0.910, 0.939]  0.939 |
| **Negative Predictive Value**  PPMI Full (Internal)  PPMI Bootstrap**  S4 (External) | 0.693  0.697 [0.662, 0.732]  0.870 | 0.720  0.721 [0.688, 0.753]  0.913 | 0.733  0.734 [0.700, 0.767]  0.909 |
| **Accuracy**  PPMI Full (Internal)  PPMI Bootstrap**  S4 (External) | 0.832  0.833 [0.813, 0.853]  0.915 | 0.851  0.850 [0.829, 0.871]  0.944 | 0.858  0.859 [0.838, 0.879]  0.930 |
| *Performance metrics were calculated based on the specific Youden Index Cut-Off for each model;  **Mean of bootstrap and 95% bootstrap percentile intervals | | | |

| **Appendix Table 6. Model performance metrics among manifest PD participants to discriminate a-Syn SAA+ vs. SAA- status using UPSIT percent correct** | | | |
| --- | --- | --- | --- |
| *Performance Metric and Dataset | Model (Youden Index Cut-Off) | | |
|  | UPSIT% Correct  (Cut-Off: 0.600) | UPSIT% Correct + Age + SCOPA5  (Cut-Off: 0.610) | UPSIT% Correct + Age + SCOPA5 + LRRK2/GBA  (Cut-Off: 0.600) |
| **Specificity**  PPMI (Internal)  S4 (External) | 0.707  0.250 | 0.756  0.500 | 0.813  0.500 |
| **Sensitivity**  PPMI (Internal)  S4 (External) | 0.848  0.935 | 0.864  0.957 | 0.870  0.957 |
| **Positive Predictive Value**  PPMI (Internal)  S4 (External) | 0.954  0.935 | 0.962  0.957 | 0.971  0.957 |
| **Negative Predictive Value**  PPMI (Internal)  S4 (External) | 0.392  0.250 | 0.435  0.500 | 0.463  0.500 |
| **Accuracy**  PPMI (Internal)  S4 (External) | 0.831  0.880 | 0.851  0.920 | 0.863  0.920 |
| *Performance metrics were calculated based on the specific Youden Index Cut-Off for each model | | | |
